# The relationship between clusters of multimorbidity and dementia risk: A systematic review

**DOI:** 10.1101/2025.03.04.25323241

**Authors:** Tiago Wiesner, Paula Grammatikos, Veerle van Gils, Sarah Bauermeister

## Abstract

**Aim:** Investigating the available evidence on the relationship between clusters of multimorbidity and dementia risk in adults.

**Methods:** Embase, PsycINFO, and Ovid MEDLINE were searched until the 9^th^ of February 2025. Included studies reported dementia risk or incidence in adult populations in relation to different clusters of multimorbidity. A narrative synthesis was structured according to the identified clusters across studies, their associations with dementia risk, and any moderation or stratification analyses for APOE ε4 allele carriership and C-reactive protein (CRP), among others. The Quality In Prognosis Studies (QUIPS) tool was used for quality assessment.

**Results:** Of the 870 abstracts screened, 7 were included in the final synthesis. Significant relationships between clusters of multimorbidity and an elevated risk of dementia were identified in all studies. The most consistent findings related to cardiometabolic and mental health/neuropsychiatric clusters evidencing the highest dementia risk. Other multimorbidity clusters were less well studied and results regarding dementia risk varied across studies. Moderation and stratification analyses for APOE ε4 and CRP, where available, yielded inconsistent findings.

**Conclusions:** This systematic review highlights the importance of understanding multimorbidity clusters for early identification of dementia risk and targeted treatment approaches. Further research is required to explore relationships between multimorbidity clusters and dementia risk across different ethnic groups as well as the potential moderating role of lifestyle factors.

**Systematic review (PROSPERO) registration number:** CRD42024619521

## Introduction

Dementia is one of the largest public health challenges of the 21^st^ century, globally ranked as the 7^th^ leading cause of death according to the World Health Organisation (WHO, 2024). The prevalence of dementia is closely associated with increased ageing, with 55 million people currently affected – a number projected to rise to 130 million by 2050 (WHO, 2023; Winblad et al., 2016). The devastating impact of dementia extends beyond the quality of life of affected individuals and their families, creating a significant economic burden that poses challenges to global societies and healthcare systems (Skaria, 2022).

Chronic conditions that have been consistently associated with an increased risk of dementia, such as depression and hypertension (Fernández Fernández et al., 2024; Kennelly & Lawlor, 2009), are frequently studied in isolation. However, as chronic conditions rarely isolated occurrences in older adults, there is need for a more holistic approach. Approximately 90% of the population aged 60 years or older suffer from two or more chronic conditions (i.e., multimorbidity), and the risk of dementia increases with each additional chronic condition (Ben Hassen et al., 2022; Calderón-Larrañaga et al., 2017; Chen et al., 2023). Multimorbidity may contribute to dementia risk due to its profound impact on individuals’ brain health. Multimorbidity has been shown to accelerate structural brain changes involving neurodegeneration and cerebrovascular pathology (Valletta et al., 2024). In addition, multimorbidity has been linked to DNA damage and dysregulated energy metabolism, particularly mitochondrial energy dysfunction (Tomkova et al., 2024), both of which are hallmark features of biological ageing. Studies also revealed that multimorbidity significantly diminishes quality of life (Makovski et al., 2019) and elevates perceived stress by limiting the ability to participate in daily activities (Stubbs et al., 2018). Furthermore, managing multiple chronic conditions requires frequent and time-consuming medical care, which may disrupt social connections and increase feelings of loneliness (Schübbe et al., 2023). Both high stress and feelings of loneliness have been consistently shown to contribute to dementia risk (Luo et al., 2023; Sutin et al., 2020). A prospective cohort study by Ben Hassen and colleagues (2022) found that baseline multimorbidity significantly increased the risk of developing dementia, with the risk rising further for individuals with three or more chronic conditions compared to those with two. Numeric measures of morbidity counts are commonly used in research (Stirland et al., 2020) but may offer limited perspectives into the complex nature of multimorbidity. Instead, investigating the *clustering* of chronic conditions may prove more meaningful. Clusters of multimorbidity (i.e., the discrete grouping of chronic conditions) have been consistently replicated across different clustering methods (Busija et al., 2019), suggesting a non-random connection between disease co-occurrences. This consistency may reflect shared biological pathways and common risk factors underlying chronic conditions (Marengoni et al., 2020). In this context, Grande et al. (2021) were the first to examine multimorbidity clusters, demonstrating that certain clusters significantly increase the risk of developing dementia. These findings underscore the potential role of interconnected disease mechanisms in driving dementia risk.

As such, understanding multimorbidity clusters may be crucial for identifying groups of people at high risk for developing dementia, thereby improving targeted prevention, intervention, and treatment strategies. The aim of this systematic review is to explore the current state of research on the associations between clusters of multimorbidity and dementia risk, exploring recurring clusters that may be critical. By conducting a narrative synthesis of the available evidence, we provide an overview of the field, identify key gaps and limitations in the literature, and offer valuable insights to guide future studies.

## Methods

### Eligibility criteria

The initial search was conducted on the 12^th^ of December 2024 and due to the rapidly growing field of multimorbidity clusters and dementia risk, a subsequent search was executed on the 9^th^ of February 2025 to update the systematic review. We did not limit the publication date. The inclusion criteria were as follows: studies published in English language, using adult populations, using multimorbidity cluster analyses as predictor, and having dementia risk or incidence as primary or secondary outcome. Studies were required to be original research articles published in peer-reviewed journals with longitudinal study designs. Studies were excluded when including individuals with a dementia diagnosis at baseline, individuals with fewer than two morbidities, qualitative studies, reviews, case reports, case series, conference abstracts, book chapters, editorials, and letters. The protocol was registered in the PROSPERO database (CRD42024619521).

### Search strategy

For our systematic review, we conducted a search across 3 databases: Embase, PsycINFO, and Ovid MEDLINE. The search terms for multimorbidity were derived from a preliminary literature review, with an emphasis on being as inclusive as possible by using multiple definitions for multimorbidity, given the inconsistency in terminology across the literature (Dunn et al., 2022). We applied standardised indexing terms, such as MeSH terms (Ovid MEDLINE), Emtree (Embase), and the APA Thesaurus (PsychInfo), to ensure a comprehensive search (see Supplementary Table S1). The search was executed in Ovid, and after removing duplicates, further screening for eligibility was completed by using Rayyan, a tool designed to conduct systematic reviews (Ouzzani et al., 2016). Additionally, the study complied with the Preferred Reporting Items for Systematic Review and Meta-analysis (PRISMA; Page et al., 2021) 2020 27-item checklist.

### Study selection

Titles, abstracts, and subsequently full-text review was conducted by two independent reviewers (TW and PG). To resolve disagreements, consensus was reached with the involvement of a third reviewer (SB).

### Data extraction and synthesis

The following information was extracted from the included studies: details of the study (author, year of publication, country, study design, sampling approach), population (sample size, sample characteristics), statistical methods used, covariates included, length of follow-up, and results (risk/incidence of dementia). All studies included in the systematic review were summarised in a narrative synthesis. Information obtained from data extraction is illustrated in Table 1 and 2.

**Table 1.** Study Design and Participant Demographics.

Study Design and Participant Demographics
| Author, year | Country | Study design | Data source | Sampling approach | Sample Size (n) | Age, y | Sex (% female) | Follow-up (y) |
| --- | --- | --- | --- | --- | --- | --- | --- | --- |
| Calvin et al., 2022 | United Kingdom | PC | UK Biobank | Adults aged at least 60 years without dementia at baseline | 206,960 | 64.1 (SD: $\pm 2.9$) | 52.7% | 15 |
| Grande et al., 2021 | Sweden | PC | SNAC-K | Adults aged at least 60 years without dementia and with multimorbidity at baseline | 2,622 | 75 (SD: $\pm 10$) | 64.3 % | 12 |
| Hu et al., 2022 | United Kingdom | PC | UK Biobank | Adults aged at least 55 years without dementia at baseline | 245,483 | 62.32 (SD: $\pm 4.08$) | 53.16% | 9 |
| Khondoker et al., 2023 | United Kingdom | PC | UK Biobank | Adults without dementia at baseline or one year from the date of recruitment | 447,888 | 58.0 (IQR: 50.0-63.0) | 54.3% | 11 |
| Patel et al., 2024 | United Kingdom | PC | UK Biobank | Adults aged at least 65 years without dementia and with at least one chronic condition at baseline | 282,712 | 61.6 (SD: $\pm 4.65$) | 53.3% | 12 |
| Valletta et al., 2023 | Sweden | PC | SNAC-K | Adults aged at least 60 years without dementia and with multimorbidity at baseline | 2,693 | 74.9 (SD: $\pm 10.5$) | 64.9% | 18 |
| Wang et al., 2024 | China | PC | CLHLS | Adults without dementia until 65 years | 14,093 | n.a. | 52.7% | 6 |

*Note.* CLHLS, Chinese Longitudinal Healthy Longevity Study; PC, prospective cohort; SNAC-K, Swedish National Study on Aging and Care in Kungsholmen.

### Quality assessment

For each study, two independent reviewers (TW and PG) rated the risk of bias based on the Quality In Prognosis Studies (QUIPS; Hayden et al., 2013), with disagreements resolved in discussion with a third reviewer (SB). The QUIPS tool evaluates potential biases across six domains: study participation, study attrition, measurement of prognostic factors, outcome measurement, study confounding, and statistical analysis and reporting. The risk of bias for each area was classified as low, moderate, or high.

### Results Study selection and cohort characteristics

Based on the initial search criteria, seven studies were included in the systematic review (see Figure 1 for PRISMA flow chart). The analysed cohorts represented three different countries (see Table 1), using the same national cohort data within each country: China (Chinese Longitudinal Health Longevity Study [CLHLS] cohort, one study), United Kingdom (UK Biobank cohort, four studies), and Sweden (Swedish National study on Aging and Care in Kungsholmen [SNAC-K] cohort, two studies). Cohort sizes ranged from 2,622 to 447,888 participants, the age in each cohort ranged from 58 (IQR: 59.0-63.0) to 75 (SD: ± 10), and between 52.7-64.9% of participants were female.

**Figure 1.**
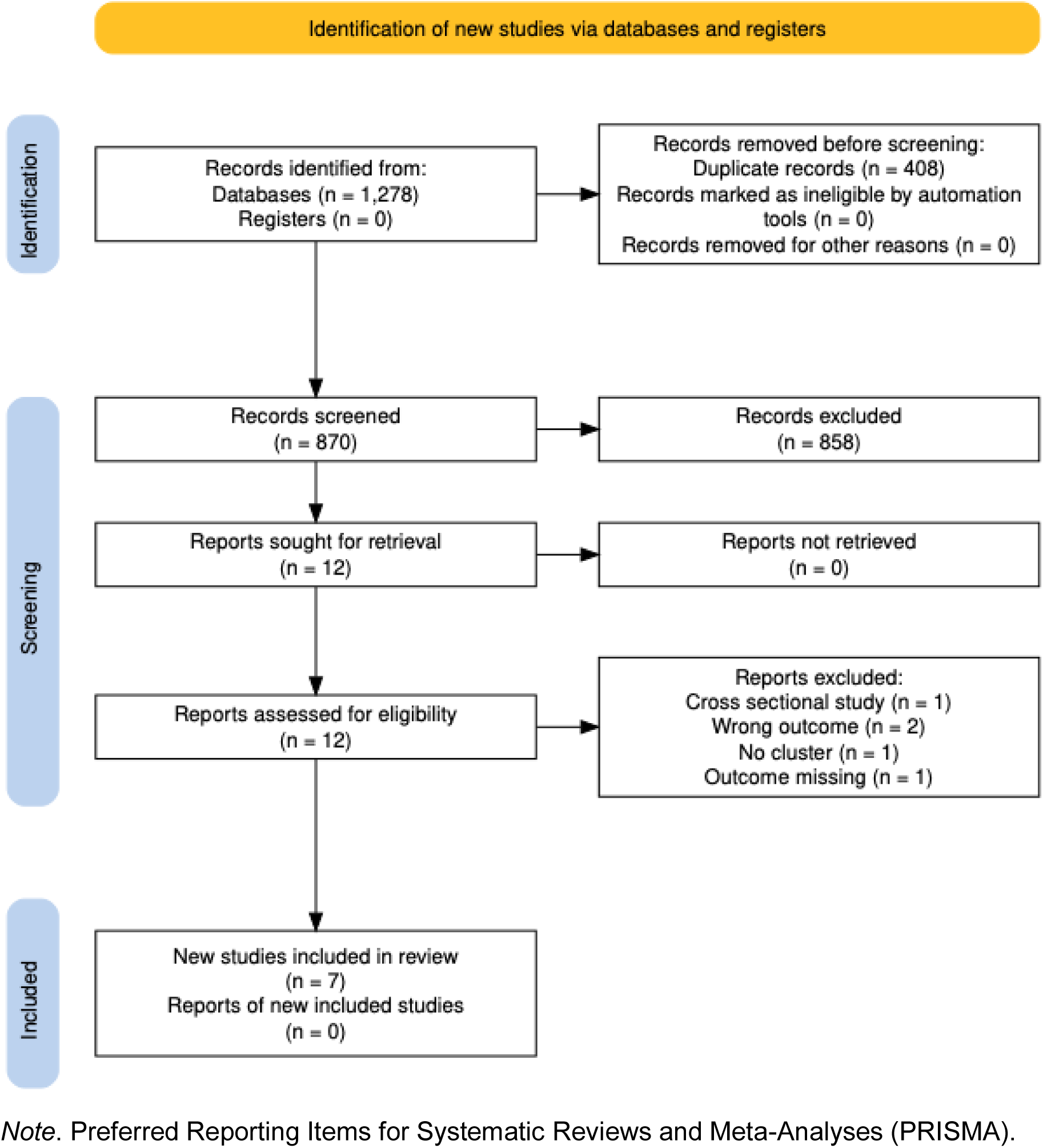
PRISMA Flowchart for the Selection of Studies

### Assessment of conditions and diversity of clusters

Between 14 and 59 conditions were included in the analyses (see Supplementary Table S2), based on the information available within cohort data and the authors’ theoretical framework. All studies defined conditions based on a combination of various ICD-10 codes, which were assessed through multiple sources, including hospital inpatient records, outpatient care data, physical examinations, and self-reports (Calvin et al., 2022; Grande et al., 2021; Hu et al., 2022; Khondoker et al., 2023; Patel et al., 2024; Valletta et al., 2023; Wang et al., 2024; see Table 2). Additionally, four distinct statistical methods (i.e., exploratory factor analysis, latent class analysis, fuzzy c-means cluster analysis, and k-means cluster analysis) were applied in order to cluster conditions (see Figure 2).

**Figure 2.**
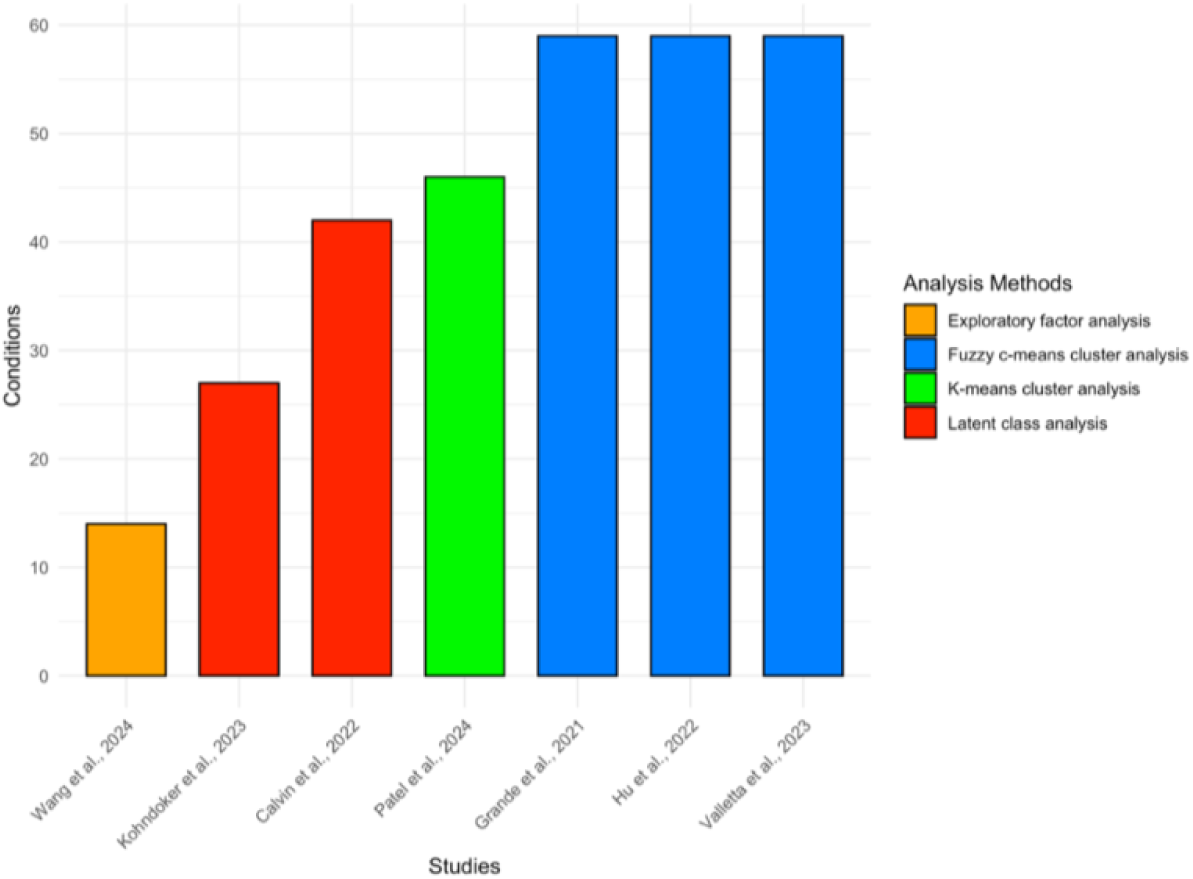
Number of Chronic Conditions by Studies and Analysis Method used

**Table 2.**
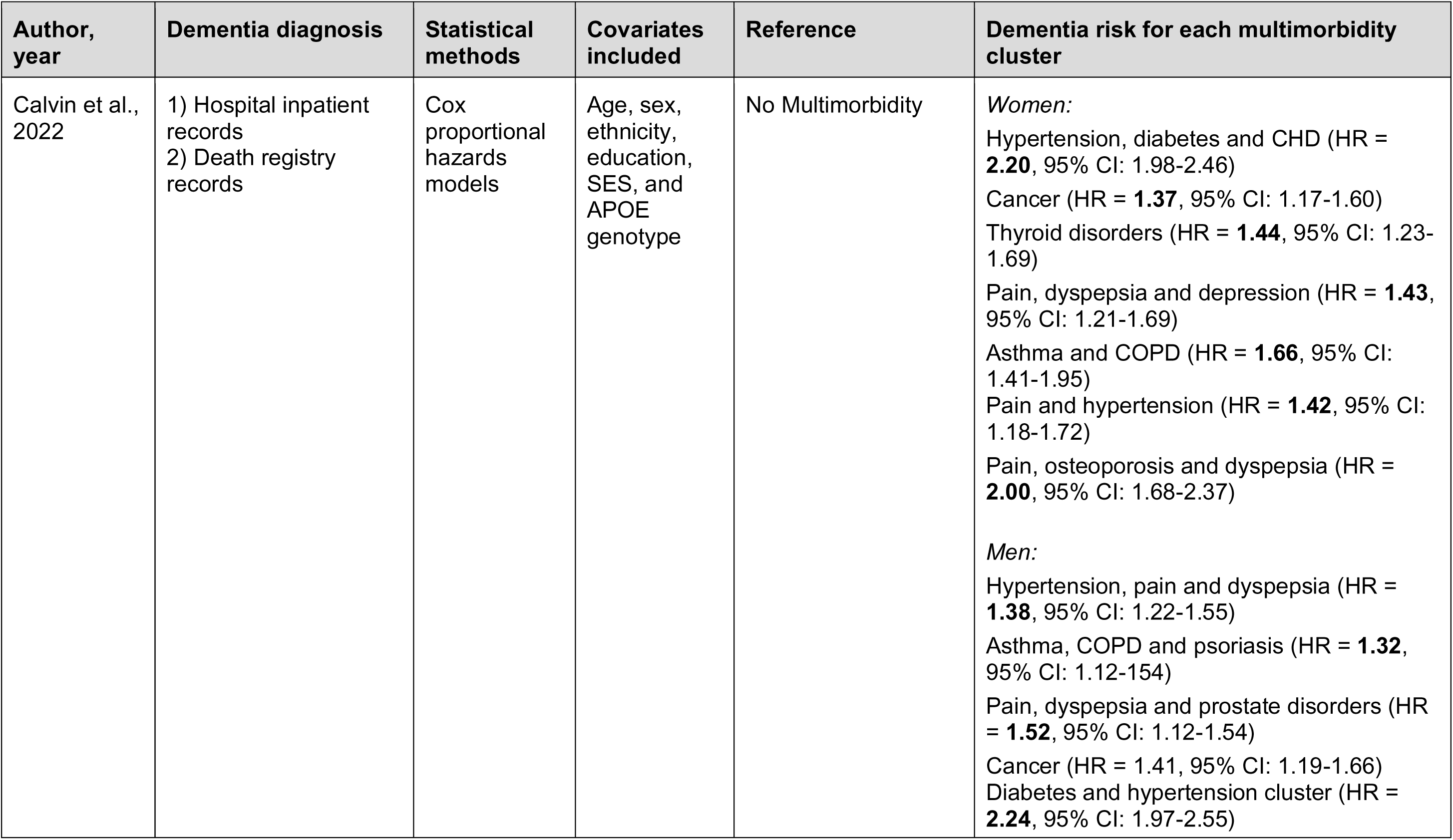
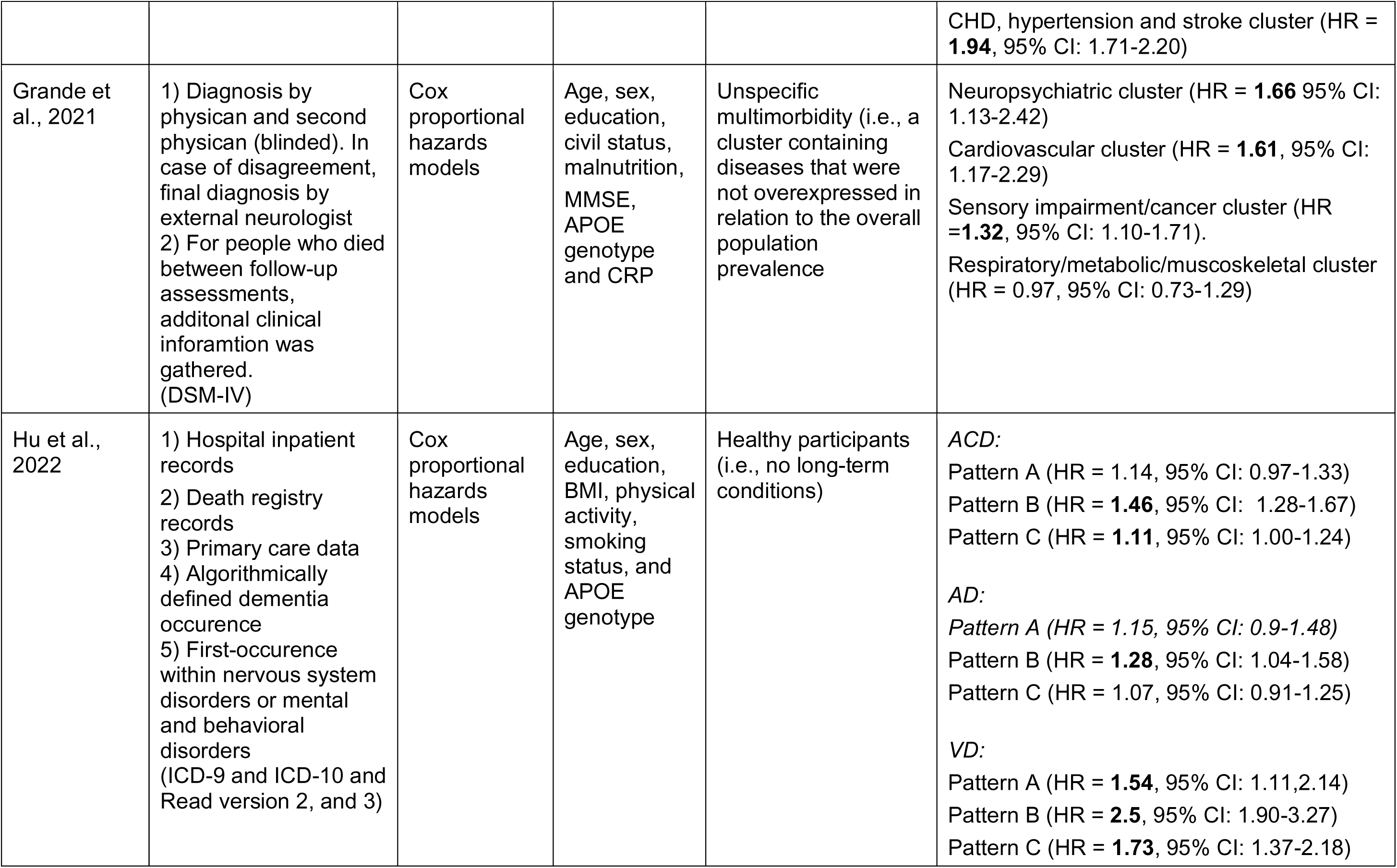
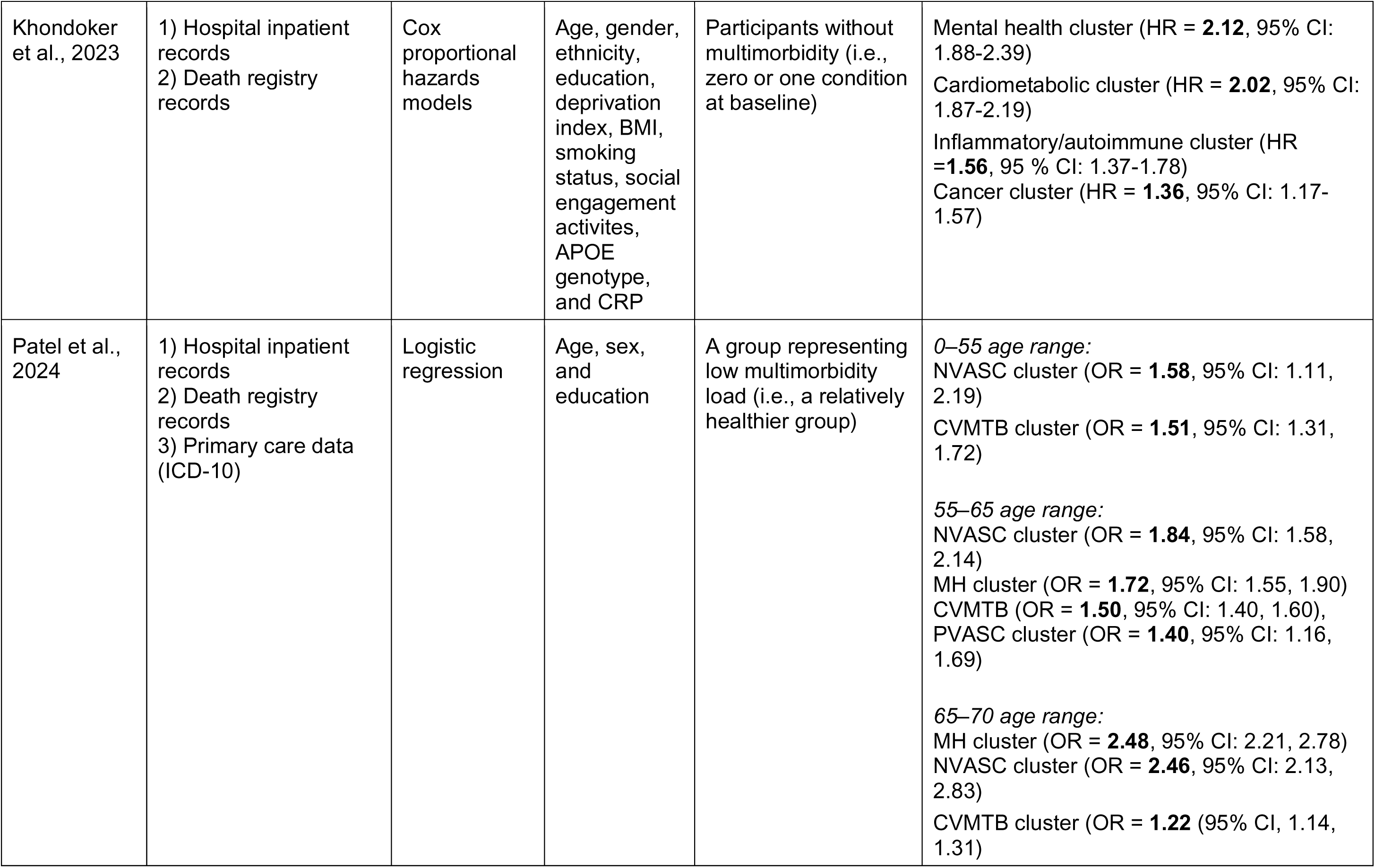
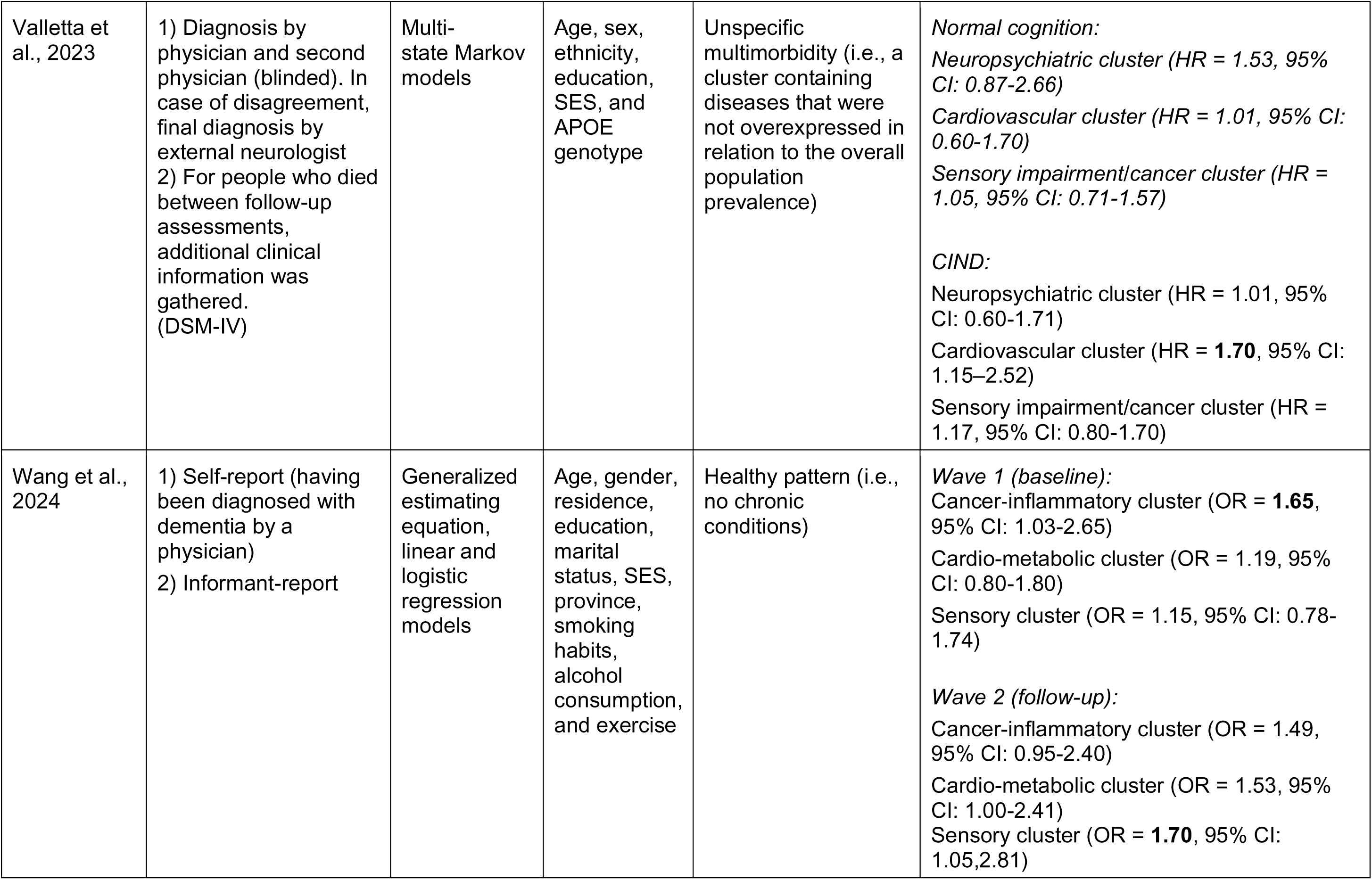
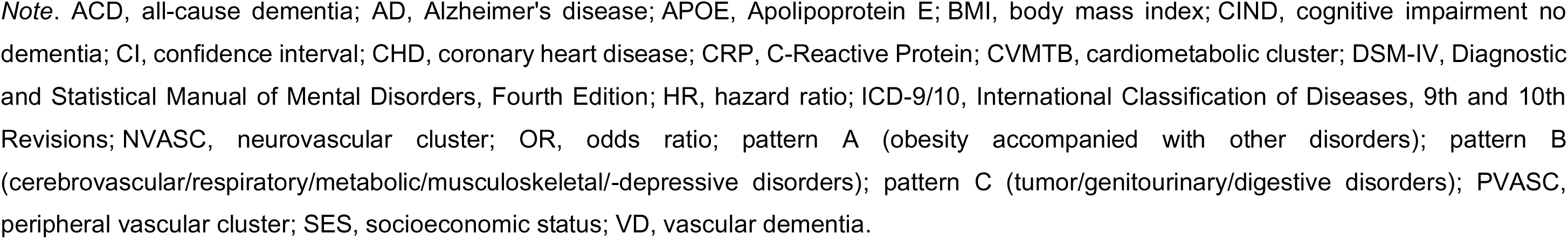
Dementia Risk Analysis: Diagnosis, Methods, and Covariates.

The defined comorbidity clusters demonstrated notable overlap across studies. Cardiometabolic clusters were identified in six out of seven studies (Calvin et al., 2022; Grande et al., 2021; Khondoker et al., 2023; Patel et al., 2024; Valletta et al., 2023; Wang et al., 2024, while cancer-related clusters appeared in five studies (Calvin et al., 2022; Grande et al., 2021; Khondoker et al., 2023; Valletta et al., 2023; Wang et al., 2024). Five studies highlighted clusters associated with mental health/neuropsychiatric conditions (Calvin et al., 2022; Grande et al., 2021; Khondoker et al., 2023; Patel et al., 2024; Valletta et al., 2023). Sensory-related clusters were observed in four studies (Grande et al., 2021; Patel et al., 2024; Valletta et al., 2023; Wang et al., 2024), respiratory clusters in three studies (Calvin et al., 2022; Grande et al., 2021; Valletta et al., 2023), and musculoskeletal-related clusters in two studies (Grande et al., 2021; Valletta et al., 2023). Additionally, inflammation-related clusters were identified in two studies (Khondoker et al., 2023; Wang et al., 2024). Some clusters were highly heterogeneous in terms of the conditions they encompassed. For instance, Hu and colleagues (2022) identified a complex cluster comprising cardio-cerebrovascular, respiratory, metabolic, musculoskeletal, and depressive disorders, while Grande et al. (2021) and Valletta et al. (2023) included a sensory impairment/cancer cluster. Patel et al. (2024) introduced more detail within cardiometabolic clusters by distinguishing between cardiometabolic, peripheral vascular, and neurovascular clusters. These examples illustrate the complexity and diversity of multimorbidity, highlighting how conditions may often co-occur in intricate patterns. For a more detailed overview, see Supplementary Table S2.

### Clusters associated with dementia

In all studies reviewed, different clusters of multimorbidity were associated with an elevated risk of developing dementia. Cardiometabolic-related clusters were most consistently linked to an increased dementia risk, as evidenced in five out of six studies (Calvin et al., 2022; Grande et al., 2021; Khondoker et al., 2023; Patel et al., 2024; Valletta et al., 2023). The strength of the association between cardiometabolic clusters and dementia varied across studies. Grande et al. (2021) reported that the cardiometabolic cluster significantly increases subsequent risk of developing dementia, with a hazard ratio (HR) of 1.61 (95% confidence interval [CI]: 1.17-2.29). Valletta et al. (2023) found that participants with cognitive impairment no dementia (CIND) within the cardiometabolic cluster had a HR of 1.70 (95% CI: 1.15–2.52). The risk of developing dementia in the cardiometabolic cluster of Khondoker et al. (2023) was higher yet, with a HR of 2.02 (95% CI: 1.87-2.19). Calvin et al. (2022), in the only study examining clusters across sexes, identified an even higher dementia risk associated with cardiometabolic-related clusters (HR of 2.20 [95% CI: 1.98-2.46] in women and 2.24 [95% CI: 1.97-2.55] in men). Finally, Patel et al. (2024) were the first to investigate the change of dementia risk and multimorbidity clusters throughout the lifespan. They calculated the odds ratios (ORs) for dementia incidence by age intervals (0-55, 55-65, 65-70), representing the odds of developing dementia in each interval. Among cardiometabolic-related clusters, an OR of 1.22 (95% CI: 1.14-1.31) was observed for the cardiometabolic cluster in the 65-70 age group, and an OR of 2.46 (95% CI: 2.13-2.83) was noted for the neurovascular cluster in the same age group, indicating a marked increase in dementia risk in both cases.

Out of the five studies that included a mental health/neuropsychiatric-related cluster, four found a significant association with dementia incidence (Calvin et al., 2022; Grande et al., 2021; Khondoker et al., 2023; Patel et al., 2024). Grande et al. (2021) identified a neuropsychiatric cluster, Calvin et al. (2022) a pain, dyspepsia, and depression cluster in women, and both Khondoker et al. (2023) and Patel et al. (2024) described a mental health cluster. The reported HRs were 1.66 (95% CI: 1.13-2.42), 1.43 (95% CI: 1.21-1.69), and 2.21 (95% CI: 1.88-2.39) for the first three, respectively. For Patel et al. (2024), the ORs differed by age group, with an OR of 1.72 (95% CI: 1.55, 1.90) for the 55-65 age group and an OR of 2.48 (95% CI: 2.21, 2.78) for the 65-70 age group.

Furthermore, cancer and sensory-related clusters have been associated with an increased risk of dementia in five out of six studies (Calvin et al., 2022; Grande et al., Khondoker et al., 2023; Valletta et al., 2023; Wang et al., 2024). Wang et al. (2024) identified an OR of 1.70 (95% CI: 1.05-2.81) for the sensory cluster and an OR of 1.65 (95% CI: 1.03- 2.65) for the cancer-inflammatory cluster, indicating a significant elevation in dementia risk. Similarly, Grande et al. (2021) reported a HR of 1.32 (95% CI: 1.10-1.71) for the sensory impairment/cancer cluster, further supporting the link between sensory-related multimorbidity and dementia incidence. Additionally, Khondoker et al. (2023) found that their cancer cluster increased the risk of dementia with a HR of 1.36 (95% CI: 1.17-1.57), while Calvin et al. (2022) reported an association in women with a HR of 1.37 (95% CI: 1.17-1.60) and a HR of 1.41 (95% CI: 1.19-1.66) in men.

Finally, the difficulty in making direct comparison with the clusters/patterns investigated by Hu et al. (2022) arises due to their heterogeneity. Therefore, they were not grouped into the previously mentioned clusters related to specific groups of conditions. However, their findings provide a nuanced perspective on dementia subtypes, highlighting the risk of developing all-cause dementia (ACD), vascular dementia (VD), and Alzheimer’s disease (AD) across different clusters. Individuals in the obesity accompanied with other disorders cluster (pattern A) showed an increased risk of developing VD with a HR of 1.54 (95% CI: 1.11-2.14), whereas the cerebrovascular/respiratory/metabolic/musculoskeletal or depressive cluster (pattern B) demonstrated an elevated risk of developing ACD with a HR of 1.46 (95% CI: 1.28- 1.67), AD with a HR of 1.28 (95% CI: 1.04-1.58), and VD with a HR of 2.50 (95% CI: 1.90-3.27). In comparison, those in the tumor/genitourinary or digestive disorder cluster (pattern C) showed a modestly increased risk for ACD with a HR of 1.11 (95% CI: 1.00-1.24) and a significantly higher risk for VD with a HR of 1.73 (95% CI: 1.37-2.18).

## Moderation and stratified analyses

### Apolipoprotein E gene variant

Khondoker et al. (2021), Grande et al. (2021), and Calvin et al. (2022) all examined the moderating effects of Apolipoprotein E gene variant (APOE ε4) status in multimorbidity clusters on dementia risk. Only the latter found that multiple multimorbidity clusters for both women and men significantly interacted with APOE ε4 status. Their results showed that the risk of developing dementia was elevated in noncarriers compared to carriers. Grande et al. (2021) extended their analysis by stratifying groups based on APOE ε4. Carriers compared to non-carriers showed a stronger association of the neuropsychiatric cluster (HR=2.79, 95% CI: 1.43-5.43) and the cardiovascular cluster (HR=2.58, 95% CI: 1.39-4.76) with dementia. In addition, Hu et al. (2022) also observed differences in the associations between multimorbidity patterns and dementia based on APOE ε4 status. Carriers demonstrated stronger associations in pattern C in ACD (HR = 1.18, 95% CI: 1.03-1.36) and pattern A in VD (HR = 1.9, 95% CI:1.17-3.08). Conversely, an amplified association has been shown in pattern B for non-carriers (HR = 1.42, 95% CI: 1,03-1.97).

### C-reactive protein

Similarly, Khondoker et al. (2023) and Grande et al. (2021) did not find any significant results when examining the moderating effects of C-reactive protein (CRP) in multimorbidity clusters on dementia risk. When stratifying groups based on CRP levels, Grande et al. (2021) observed significant differences across the various clusters. The association between neuropsychiatric, cardiovascular, and sensory impairment/cancer clusters and dementia was notably stronger in the group with high CRP levels, with HRs of 2.69 (95% CI: 1.19-6.09), 2.34 (95% CI: 1.08-5.09), and 1.92 (95% CI: 1.02–3.63), respectively.

### Sex and age

Hu et al. (2022) stratified their analyses by sex, revealing that females exhibited an increased risk of ACD with a HR of 1.25 (95% CI:1.06-1.47) for pattern C compared to males, whereas males showed a higher risk of VD for pattern A (HR = 1.72, 95% CI: 1.14-2.6) and C (HR = 1.91, 95% CI: 1.42-2.57) compared to females. Additionally, they stratified by age, but did not find any differences.

### Quality assessment

Based on the evaluation with the QUIPS tool, no study was excluded as the general risk of bias was low across all studies. However, most studies exhibited a moderate to high risk of bias on the study attrition domain, and a low to moderate risk in the outcome measurement domain. A summary can be found in Supplementary Table S3.

## Discussion

Accumulated findings from this systematic review consistently demonstrate associations between specific multimorbidity clusters and an increased risk of developing dementia. The most promisingly clusters that are associated with the risk of dementia in this review were cardiometabolic and mental health/neuropsychiatric clusters. This underscores the importance of considering the complex and interconnected nature of multimorbidity rather than focusing on multimorbidity counts or isolated conditions.

### Potential linkage of co-occurring conditions

Cardiometabolic conditions, for instance stroke, hypertension, diabetes, and high levels of low-density lipoprotein (LDL) cholesterol, are well-established risk factors of dementia (Livingston et al., 2024). Recent findings suggest that an *accumulation* of multiple cardiometabolic conditions (i.e., heart disease, stroke, and diabetes) substantially increase the risk of dementia (Dove et al., 2023; Tai et al., 2022). Further reinforcing these findings, all but one study in this review significantly associated cardiometabolic-related multimorbidity with an increased risk of developing dementia. Cardiometabolic conditions often share common underlying pathologies such as atherosclerosis (Santos et al., 2017), which has been associated with AD and increased risk of cognitive decline (Xie et al., 2020). Similarly, chronic hypertension leads to vascular changes including hypertrophy, which, like atherosclerosis, may reduce cerebral blood flow (Han et al., 2019; Vaudo et al., 2000). This decline in cerebral blood flow contributes to hypoxia, amyloid β (Aβ) protein accumulation, and blood brain barrier disruption (Duncombe et al., 2017). These changes increase neuronal dysfunction, highlighting their impact in the pathophysiology of dementia, particularly vascular dementia. Another crucial factor implicated in cardiometabolic conditions, dementia, and vascular brain damage is inflammation (Darweesh et al., 2018; Donath et al., 2019; Mun & Hinman, 2022). Darweesh and colleagues (2018) identified CRP as one of the inflammatory markers associated with all-cause dementia. Interestingly, Khondoker et al. (2023) and Grande et al. (2021) both examined the moderating effects of CRP in multimorbidity clusters, but neither study found significant results. Grande et al. (2021) subsequently argue for investigating the mediating role of CRP in the relationship between multimorbidity and dementia risk, whereas Khondoker et al. (2023) instead suggest that CRP influences dementia through its effect on multimorbidity. Grande et al. (2021) demonstrated, by stratifying clusters for CRP, that high CRP levels amplify the risk of developing dementia in all clusters, except the respiratory/metabolic/musculoskeletal cluster, indicating that the impact of inflammation on dementia may differ between multimorbidity clusters. The precise role of CRP in multimorbidity-related dementia risk thus remains unresolved. Future research may also benefit from incorporating additional inflammatory markers.

In addition to cardiometabolic conditions, mental health/neuropsychiatric-related clusters were also consistently associated with an increased risk for dementia. The most frequent and dominant conditions in these clusters were depression, anxiety, schizophrenia, and stress-related conditions such as post-traumatic stress disorder (PTSD). Common biological dysregulations, including the HPA axis, neurotransmission, and elevated inflammation, are implicated in these conditions (Lawrence & Scofield, 2024; Michopoulos et al., 2017; Mikulska et al., 2021). These dysregulations may initiate processes that increase the brain’s vulnerability to neurodegeneration and cognitive impairment. Pro-inflammatory cytokines, for example, stimulate the production of Aβ and hyperphosphorylated tau protein, and vice versa, creating a vicious cycle that leads to neuronal damage (Novoa et al., 2022). Furthermore, research has shown that alterations in neurotransmitter systems may cause neurotoxic effects leading to neurodegeneration and, ultimately, dementia (Jha et al., 2017). Additionally, an emerging and strongly debated factor in the aetiology of mental health conditions is the gut microbiome (i.e., microorganisms in the gastrointestinal tract; Butler et al., 2019). Calvin and colleagues (2022) identified a cluster of dyspepsia, one of the most researched chronic gastrointestinal disorders (Sperber et al., 2021), depression, and pain. Interestingly, dyspepsia may be associated with an alteration of the function and composition of intestinal microbes impacting the microbiome (Brown et al., 2022), which may help explain the observed co-occurrence of these conditions. Notably, the risk of dementia was increased within this cluster, highlighting the potential impact of microbiome dysbiosis on dementia.

Two studies in this review have illustrated the potential risk of dementia associated with clusters related to sensory impairments (Grande et al., 2021; Valletta et al., 2023). A strong consensus has developed regarding sensory impairments, such as hearing loss, and their risk for developing dementia (Bucholc et al., 2021; Griffiths et al., 2020; Loughrey et al., 2018). Griffiths et al. (2020) point out that hearing loss may independently increase dementia risk by reducing cognitive reserve through a deprived auditory environment, leading to negative effects on brain structure and function. Additionally, impaired speech perception renders social interactions difficult, which may further contribute to dementia development.

Clusters of cancer incidence were also identified. Evidence regarding cancer and its potential association with dementia is rather mixed. Whereas some studies demonstrate that there might be an inverse relation between cancer survivors and dementia risk (Yarchoan et al., 2017; Zhang et al., 2022), other studies found that cancer survivors are more likely to develop dementia (e.g., Chu et al., 2025).

Notably, Grande et al. (2021) even identified a combined sensory impairment/cancer cluster that was associated with dementia. A possible explanation for this finding may be that chemotherapy, which is being used to combat cancer, may not only lead to chemotherapy-induced cognitive impairment (Ahles et al., 2012), but also fuel sensory impairments such as hearing loss (Frisina et al., 2016), driving the potential link between cancer and sensory conditions. Interestingly, chronic conditions, such as hearing loss, hypertension and depression are also captured by recent developed dementia risk scores (Rosenau et al., 2024), reflecting their importance in the realm of dementia research. Shang et al. (2022) even created a multimorbidity risk score for dementia incorporating age and APOE ε4. Their findings suggest that multimorbidity may have a greater impact on dementia risk in non-carriers, potentially overshadowing genetic effects. Supporting this, Calvin et al. (2022) also reported that non-carriers were, in relative terms, at an elevated risk compared to carriers. Although the role of APOE ε4 role in moderating the effect of clusters of multimorbidity on dementia remains inconsistent (Calvin et al., Grande et al., 2021; Khondoker et al., 2023), research suggests that preventing multimorbidity could help reduce dementia risk, particularly in non-carriers.

### Available data and applied statistical methods

Differences in the clusters and outcome of dementia risk may, of course, depend to some extent on the available data of the cohorts. In this review, four studies were based on the UK Biobank and two were based on the Swedish SNAC-K cohort. The mean age of the UK Biobank ranged between 58 and 64 (Calvin et al., 2022; Hu et al., 2022; Khondoker et al., 2023 Patel et al., 2024), whereas the mean age for the SNAC-K studies was approximately 75 (Grande et al., 2021; Valletta et al., 2023). Interestingly, Patel et al. (2024) found not only that the risk of multimorbidity clusters on dementia varies within different age groups, but also that cluster definitions change depending on age. Therefore, the difference in age needs to be considered when comparing results across different cohorts, as this may have a potential impact on the specific cluster composition and their associated risk of dementia.

Beyond that, many other components, such as varying chronic conditions included and lifestyle factor profiles of participants, may differ in the respective cohorts and countries. To illustrate the difference in outcome, we can compare the results of Hu et al. (2022) and Grande et al. (2021). Both studies used similar methodological approaches. They included the same conditions (varied based on the data available) and both applied fuzzy c-means cluster analysis, but given the differences in the cohorts used, there were differences in the composition of clusters and their associated dementia risk.

Furthermore, the clustering method varied across studies (see Figure 2), which may result in potential heterogeneity in the observed clusters. This is highlighted by a systematic review of 51 studies (Busija et al., 2019), which found that clusters of multimorbidity were less replicable than others across different clustering methods. Crucially, it remains challenging to identify clinically meaningful clusters given the complexity of clusters in studies. As an example, one of the clusters in Hu et al.‘s (2022) analysis ranged from cardiocerebrovascular diseases to respiratory, metabolic, musculoskeletal, and depressive disorders, making its consideration for future clinical decisions difficult. Therefore, future studies should agree on clinically relevant criteria before identifying multimorbidity clusters (Busija et al., 2019).

### Strengths

This systematic review offers a comprehensive synthesis of all relevant studies on multimorbidity clusters and their impact on dementia risk, providing valuable insights into this emerging field. It expands on previous work (Xin et al., 2025) by integrating recent evidence and shedding new light on the complex relationship between multimorbidity and dementia risk. We thoroughly examined moderation and stratified analyses for APOE genotype and CRP in the association between multimorbidity clusters and dementia and explored the various methodologies used to define and analyse these clusters. The present review helps to paint a clearer picture of the complex differences between studies and provides concrete guidance for future research designs and analyses.

## Limitations and future directions

A number of limitations need to be addressed. Even though some overlap between defined multimorbidity clusters exists, the heterogeneity of clusters renders their comparability complex. Additionally, only three cohorts were assessed across the seven studies, and six of them primarily represented two high-income countries (United Kingdom & Sweden), thus decreasing generalisability. In line with this, cohorts included in this review were predominantly White. Eto et al. (2023) found that clusters of multimorbidity vary across different ethnicities in the UK. Therefore, future investigations should focus on low- and middle-income countries and explore clusters in non-white ethnicities/ethnic minorities in high-income countries. Moreover, polypharmacy, which was not accounted for in these studies, is a key aspect of multimorbidity. The number and type of medications taken could provide valuable insights for future research on multimorbidity clusters and their association with dementia risk. In addition, the severity and duration of chronic conditions were not assessed in this review, which may have influenced the findings. Since most studies collected data on chronic conditions through self-reports, the results may also be affected by recall bias and classification inaccuracies. Some studies have analysed moderating effects of CRP and APOE ε4, but the role of lifestyle factors in this context remains largely unexplored. Future research could benefit from examining lifestyle factors, as they may play a key role especially in cardiometabolic clusters. Crucially, mixed findings regarding cognitive outcomes were present in different lifestyle interventions for dementia (Andrieu et al., 2017; Ngandu et al., 2015; Sakurai et al., 2024; Zülke et al., 2024). Dove et al. (2024), for instance, found that an anti-inflammatory diet was associated with a lower dementia risk and reduced neurodegenerative and vascular brain damage in individuals with cardiometabolic diseases. Therefore, accounting for an individual’s specific multimorbidity profile, lifestyle interventions could become more personalised, addressing the unique combination of health conditions that contribute to dementia risk.

## Conclusion

The growing shift in clinical practice toward a more holistic approach—considering multiple coexisting diseases rather than treating conditions in isolation—highlights the importance of researching multimorbidity clusters. This review underscores the consistent association between multimorbidity clusters, in particular cardiometabolic-related and mental health/neuropsychiatric-related clusters, and dementia risk, emphasising the need for further investigation in this emerging field. A deeper understanding of these clusters could enable more targeted treatments, interventions, and prevention strategies, ultimately reducing the projected rise in incidences of dementia in the future.

## Data Availability

All data produced in the present work are contained in the manuscript.

