## Supplementary Tables S1, S2 and S3 for "The relationship between clusters of multimorbidity and dementia risk: A systematic review"

**Supplementary Materials**

**Table S1**

*Electronic Literature Search of Embase, Ovid MEDLINE, and PsycINFO through Ovid*

| Database | Steps | Search Terms |
|---|---|---|
| Embase, Ovid MEDLINE, PsycINFO | 1 | exp multimorbidity/** or exp comorbidity/ or exp "multiple chronic conditions"/** or multimorbidit* or comorbidit* or co-morbidit* or polymorbidit* or "patient complexit*" |
| | 2 | exp dementia/ or dementia or alzheimer* disease or AD |
| | 3 | exp Risk/** or exp Incidence/** or inciden* or risk* |
| | 4 | Exp Cluster Analysis/ or cluster* or cluster analys* or pattern* or factor analys* |
| | 5 | 1 and 2 and 3 and 4 |
| | 6 | 5 not (exp qualitative research/ or exp case report/ or exp case series/ or exp review/ or exp conference abstract/ or exp book chapter/ or exp editorial/ or exp letter/) |
| | 7 | limit 6 to (adult <18 to 64 years> or aged <65+ years>) *** |
| | 8 | limit 7 to English language |
| | 9 | remove duplicates from 8 |

*Note.* Different definitions of morbidity and comorbidity were identified through a scoping review (Dunn et al., 2022), highlighting the heterogeneity in how these concepts are operationalized. A comprehensive search was conducted using the Ovid interface to access Embase, Ovid MEDLINE, and PsycINFO databases. To ensure an inclusive and exhaustive search strategy, the term finder function was utilized to identify and incorporate relevant Emtree terms (specific to Embase), MeSH terms (specific to Ovid MEDLINE), and Thesaurus terms (specific to PsycINFO). *truncation. **the subject heading is invalid in APA PsycInfo. ***only works for Embase. Additional studies that are not eligible were removed in Rayyan.

**Table S2**

*Conditions and Clustering Methods*

| Author, year | Conditions (n) | Conditions | Cluster method | Clusters |
|---|---|---|---|---|
| Calvin et al., 2022 | 42 | Alcohol problems, Anorexia or bulimia, Anxiety & other neurotic, stress-related & somatoform disorders, Asthma, Atrial fibrillation, Bronchiectasis, Cancer (any), Chronic fatigue syndrome, Chronic kidney disease, Chronic liver disease, Chronic obstructive pulmonary disease, Chronic sinusitis, Coronary heart diesease, Depression, Diabetes, Diverticular disease of intestine, Endometriosis, Epilepsy, Glaucoma, Heart failure, Hypertension, Inflammatory bowel disease, Irritable bowel syndrome, Ménière's disease, Migraine, Multiple sclerosis, Osteoporosis, Other psychoactive substance misuse, Parkinson's disease, Painful condition, Peripheral vascular disease, Pernicious anaemia, Polycystic ovary, Prostate disorders, Psoriasis or eczema, Rheumatoid arthritis, other inflammatory polyarthropathies & systemic connective tissue disorders, Schizophrenia (and related non-organic psychosis) or bipolar disorder, Stroke and TIA, Thyroid disorders, Treated constipation, Treated dyspepsia and Viral hepatitis. | Latent class analysis | Women:<br>1. Hypertension, Diabetes and CHD<br>2. Cancer<br>3. Thyroid Disorder<br>4. Depression, pain, anxiety<br>5. Pain and Hypertension<br>6. Asthma, COPD, Psorasis<br>7. Pain, Osteoporosis, Dyspepsia<br><br>Men<br>1. Hypertension, CHD, dyspepsia<br>2. Pain, hypertension<br>3. Diabetes, hypertension, CHD<br>4. Asthma, psorasis, COPD<br>5. Dyspepsia, cancer, CHD<br>6. Depression, dyspepsia, anxiety |

| | | | | |
|---|---|---|---|---|
| Grande et al., 2021 | 59 | Allergy, Anemia, Asthma, Atrial fibrillation, Autoimmune disease, Blindness and visual impairment, Blood and blood-forming organ disease, Bradycardias and conduction diseases, Cardiac valve diseases, Cataract and other lens diseases, Cerebrovascular disease, Chromosomal abnormalities, Chronic infectious diseases, Chronic kidney disease, Chronic liver diseases, Chronic pancreas, biliary tract and gallbladder diseases, Chronic ulcer of the skin, Colitis and related diseases, Chronic obstructive pulmonar diseases, chronic bronchitis and emphysema, Deafness and hearing impairment, Depression and mood diseases, Diabetes, Dorsopathies, Dyslipidemia, Ear, nose and throat diseases, Epilepsy, Esophagus, stomach and duodenum diseases, Glaucoma, Heart failure, Hematological neoplasms, Hypertension, Inflammatory arthropathies, Inflammatory bowel disease, Ischemic heart disease, Migraine and facial pain syndromes, Multiple sclerosis, Neurotic, stress-related and somatoform diseases, Obesity, Osteoarthritis and other degenerative joint diseases, Osteoporosis, Other cardiovascular diseases, Other digestive diseases, Other eye diseases, Other genitourinary diseases, Other metabolic diseases, Other musculoskeletal and joint diseases, Other neurological diseases, Other psychiatric behavioral diseases, Other respiratory diseases, Other skin diseases, Parkinson and parkinsonism, Peripheral neuropathy, Peripheral vascular disease, Prostate diseases, Schizophrenia and delusional diseases, Sleep disorder, Solid neoplasms, Thyroid diseases, Venous/lymphatic diseases. | Fuzzy c-means cluster analysis | 1. Neuropsychiatric<br>2. Cardiovascular<br>3. Repiratory/metabolic/musculoskeletal<br>4. Sensory impairment/cancer<br>5. Unspecific multimorbidity |

| | | | | |
|---|---|---|---|---|
| Hu et al., 2022 | 59 | see Grande et al., 2021 | Fuzzy c-means cluster analysis | 1. Obesity accompanied with other disorders (pattern A)<br>2. Cardio-cerebrovascular /respiratory/ metabolic/musculoskeletal/depressive disorders (pattern B)<br>3. Tumor/genitourinary/digestive disorders (pattern C) |
| Khondoker et al., 2023 | 27 | Anxiety, Asthma, Bronchiectasis, Cancer, COPD, Depression, Diabetes, Diverticulosis, Epilepsy, Glaucoma, Hearing loss, Heart, Hypercholesterolaemia, Hypertension, IBD, Kidney, Liver, Migraine, Multiple sclerosis, Parkinson's, Prostate, Psoriasis, Rheumatoid arthritis, Schizophrenia, Stroke, Thyroid, Viral hepatitis | Latent class analysis | 1. Cancer<br>2. Cardiometabolic<br>3. Inflammation<br>4. Mental Health |
| Patel et al., 2024 | 46 | Anaemia, Anxiety disorders, Arthritis, Asthma, Atherosclerosis, Atrial fibrillation, Bipolar disorder, Bronchiectasis, Cancer, Cataract, Chronic fatigue syndrome, Chronic kidney disease, Chronic obstructive pulmonary diesease, Chronic sinusitis, Connective tissue disease, Coronary heart disease, Diabetes, Depression, Diverticular disease, Endometriosis, Epilepsy, Glaucoma, Gastro-oesophageal reflux disease, Hearing loss, Heart failure, Hypercholesterolemia, Hyperlipidemia, Hypertension, Hypothyroidism, Inflammatory bowel disease, Irritable bowel syndrome, Liver disease, Lipid disorders, Macular degeneration, Ménière's disease, Migraine, Multiple sclerosis, Obesity, Osteoporosis, Parkinson's disease, Peripheral vascular disease, Prostate conditions, Psoriasis/Eczema, Psychoses, Psychiatric disorders, Primary ciliary dyskinesia, Sleep apnoea, Sleep disorders, Stress disorders, Stroke, Transient ischaemic attack, Thyroid conditions, Ulcer. | K-means cluster analysis | 1. LOW<br>2. Neurovascular<br>3. Mental Helth<br>4. Eye<br>5. CVMTB<br>6. PVASC |

| | | | | |
|---|---|---|---|---|
| Valletta et al., 2023 | 59 | see Grande et al., 2021 | Fuzzy c-means cluster analysis | 1. Neuropsychiatric<br>2. Cardiovascular<br>3. Repiratory/metabolic/musculoskeletal<br>4. sensory impairment/cancer |
| Wang et al., 2024 | 14 | Diabetes, heart disease, stroke or cerebrovascular disease, respiratory disease, tubercolosis, cancer, gastric or duodenal ulcer, Parkinsons's disease, bedsore and prostate diseases<br>Assessed:<br>Blood pressure, hypertension, vision impairment and hearing loss | Expolatory factor analysis | 1. Cancer-inflammatory<br>2. Cardiometabolic<br>3. Sensory |

*Note*. CHD, coronary heart disease; LOW, low disease burden; CVMTB, cardiometabolic cluster, PVASC, peripheral vascular cluster; COPD, chronic obstructive pulmonary disease.

**Table S3**

*Quality in Prognostic Studies (QUIPS) Tool Assessment*

| Authors | Study Participation | Study Attrition | Prognostic Factor Measurement | Outcome Measurement | Study Confounding | Statistical Analysis and Reporting |
|---|---|---|---|---|---|---|
| Calvin et al., 2022 | Low | High | Low | Moderate | Low | Low |
| Grande et al., 2021 | Low | Moderate | Low | Low | Low | Low |
| Hu et al., 2022 | Low | High | Low | Moderate | Low | Low |
| Khondoker et al., 2023 | Low | High | Low | Moderate | Low | Low |
| Patel et al., 2024 | Low | High | Low | Moderate | Moderate | Low |
| Valletta et al., 2023 | Low | High | Low | Low | Low | Low |
| Wang et al., 2024 | Low | High | Low | Moderate | Low | Low |

*Note*. Low = low risk of bias; Moderate = moderate risk of bias; High = high risk of bias.
